# Contact Tracing of COVID-19 in Karnataka, India: Superspreading and Determinants of Infectiousness and Symptomaticity

**DOI:** 10.1101/2020.12.25.20248668

**Authors:** Mohak Gupta, Giridara G Parameswaran, Manraj S Sra, Rishika Mohanta, Devarsh Patel, Amulya Gupta, Bhavik Bansal, Archisman Mazumder, Mehak Arora, Nishant Aggarwal, Tarun Bhatnagar, Jawaid Akhtar, Pankaj Pandey, Vasanthapuram Ravi, Giridhara R Babu

## Abstract

We analysed SARS-CoV-2 surveillance and contact tracing data from Karnataka, India up to 21 July 2020. We estimated metrics of infectiousness and the tendency for superspreading (overdispersion), and evaluated potential determinants of infectiousness and symptomaticity in COVID-19 cases. Among 956 cases confirmed to be forward-traced, 8.7% of index cases had 14.4% of contacts but caused 80% of all secondary cases, suggesting significant heterogeneity in individual-level transmissibility of SARS-CoV-2 which could not be explained by the degree of heterogeneity in underlying number of contacts. Secondary attack rate was 3.6% among 16715 close contacts. Transmission was higher when index case was aged >18 years, or was symptomatic (adjusted risk ratio, aRR 3.63), or was lab-confirmed ≥4 days after symptom onset (aRR 3.01). Probability of symptomatic infection increased with age, and symptomatic infectors were 8.16 times more likely to generate symptomatic secondaries. This could potentially cause a snowballing effect on infectiousness and clinical severity across transmission generations; further studies are suggested to confirm this. Mean serial interval was 5.4 days. Adding backward contact tracing and targeting control measures to curb super-spreading may be prudent. Due to low symptomaticity and infectivity, interventions aimed at children might have a relatively small impact on reducing transmission.

**Structured Abstract:** *Background:* India has experienced the second largest outbreak of COVID-19 globally, yet there is a paucity of studies analysing contact tracing data in the region. Such studies can elucidate essential transmission metrics which can help optimize disease control policies.

*Methods:* We analysed contact tracing data collected under the Integrated Disease Surveillance Programme from Karnataka, India between 9 March and 21 July 2020. We estimated metrics of disease transmission including the reproduction number (R), overdispersion (k), secondary attack rate (SAR), and serial interval. R and k were jointly estimated using a Bayesian Markov Chain Monte Carlo approach. We evaluated the effect of age and other factors on the risk of transmitting the infection, probability of asymptomatic infection, and mortality due to COVID-19.

*Findings:* Up to 21 July, we found 111 index cases that crossed the super-spreading threshold of ≥8 secondary cases. R and k were most reliably estimated at R 0.75 (95% CI, 0.62-0.91) and k 0.12 (0.11-0.15) for confirmed traced cases (n=956); and R 0.91 (0.72-1.15) and k 0.22 (0.17-0.27) from the three largest clusters (n=394). Among 956 confirmed traced cases, 8.7% of index cases had 14.4% of contacts but caused 80% of all secondary cases. Among 16715 contacts, overall SAR was 3.6% (3.4-3.9) and symptomatic cases were more infectious than asymptomatic cases (SAR 7.7% vs 2.0%; aRR 3.63 [3.04-4.34]). As compared to infectors aged 19-44 years, children were less infectious (aRR 0.21 [0.07-0.66] for 0-5 years and 0.47 [0.32-0.68] for 6-18 years). Infectors who were confirmed ≥4 days after symptom onset were associated with higher infectiousness (aRR 3.01 [2.11-4.31]). Probability of symptomatic infection increased with age, and symptomatic infectors were 8.16 (3.29-20.24) times more likely to generate symptomatic secondaries. Serial interval had a mean of 5.4 (4.4-6.4) days with a Weibull distribution. Overall case fatality rate was 2.5% (2.4-2.7) which increased with age.

*Conclusion:* We found significant heterogeneity in the individual-level transmissibility of SARS-CoV-2 which could not be explained by the degree of heterogeneity in the underlying number of contacts. To strengthen contact tracing in over-dispersed outbreaks, testing and tracing delays should be minimised, retrospective contact tracing should be considered, and contact tracing performance metrics should be utilised. Targeted measures to reduce potential superspreading events should be implemented. Interventions aimed at children might have a relatively small impact on reducing SARS-CoV-2 transmission owing to their low symptomaticity and infectivity. There is some evidence that symptomatic cases produce secondary cases that are more likely to be symptomatic themselves which may potentially cause a snowballing effect on infectiousness and clinical severity across transmission generations; further studies are needed to confirm this finding.

*Funding:* Giridhara R Babu is funded by an Intermediate Fellowship by the Wellcome Trust DBT India Alliance (Clinical and Public Health Research Fellowship); grant number: IA/CPHI/14/1/501499.

## Introduction

COVID-19, a pneumonia caused by the novel coronavirus SARS-CoV-2 originated in Wuhan, China (1). As of 18th October 2020, the disease has spread to over 200 countries and territories, causing over 39.81M cases and 1.11M deaths of which India contributed to 7.5M cases with over 100,000 deaths. (2) Karnataka, a south-Indian state inhabited by more than 61 million people, (3) confirmed its first COVID-19 case on 9th March 2020. By 18th October, Karnataka had the third-highest COVID-19 case burden of all states in India with 765,586 cumulative cases.

Contact tracing remains one of the key public health responses in infectious disease control with a history that can be traced back to the late 19th century. (4) In the present COVID-19 pandemic, contact tracing may achieve significant outbreak control when effective reproduction number is lower due to social distancing and nonpharmaceutical interventions (NPIs). (5) In addition, primary contact tracing data is extremely valuable in elucidating transmission characteristics of an infectious disease which can be used to inform policy.

A disease’s basic reproductive number R0 describes the ‘average’ number of secondary infections (offsprings) generated by a single infected individual. Super-spreader events (SSEs) highlight a major limitation of the concept of R0, which is an average and does not capture the heterogeneity of infectiousness. (6) Each infected case does not produce R0 offspring; a small number of individuals may be responsible for a large percentage of secondary infections, whereas most others infect no one. When this occurs, the offspring distribution is said to be overdispersed. The dispersion parameter, k is smaller when superspreading plays a larger role in transmission. Studying overdispersion is crucial since most of the transmission can be eliminated if events and settings conducive to superspreading can be limited by implementing targeted measures, as opposed to overarching policies that would be needed if overdispersion was low and transmission was homogeneous. (7,8)

Serial interval, defined as the interval between symptom onset of the index case and the secondary case in a transmission chain, is a key epidemiological measure that determines the spread of an infectious disease. The serial interval is an essential metric in epidemic transmission models and in estimating reproduction numbers used to evaluate the impact of interventions and to inform policy response. (9) Studies across the world estimate the serial interval of SARS-CoV-2 between 3.9-7.5 days. (10) Estimation requires high quality data from primary contact tracing which establishes linkage between transmission pairs and thus, data-backed evidence of serial interval of SARS-CoV-2 in India and other low-resource settings is extremely limited if any. Whereas the secondary attack rate which measures the risk of infection in contacts has been studied in India by Laxminarayanan et al., (11) a comprehensive study looking at superspreading, overdispersion and serial interval has not yet been done in India, mostly owing to a lack of data.

Contact tracing of COVID-19 in Karnataka was driven by multi-sectoral teams and backed by technology, enabling the state to have one of India’s most effective contact tracing systems, at least during the early epidemic. (12) Among all Indian states, Karnataka was found to have the highest quality of COVID-19 data reporting in daily bulletins released by the state government. (13) In this study, we aimed to gain insights for disease control by understanding the transmission dynamics of SARS-CoV-2 based on surveillance and tracing data from Karnataka. We estimated the reproduction number (R), overdispersion (k), secondary attack rate (SAR), and serial interval from data and reconstructed major transmission networks. We evaluated the effect of age and other factors on the probability of asymptomatic infection, risk of transmitting the infection further, and mortality due to COVID-19. We also attempt to quantify the effect of contact tracing on reducing onward transmission and on minimising testing delays.

## Methods

### Data sources

We used data generated through surveillance activities undertaken by the Integrated Disease Surveillance Program (IDSP) and the Department of Health and Family Welfare in accordance with national and state policies. Data was de-identified before extraction and analysis. We created a composite dataset by combining daily COVID-19 bulletins (14) released by the Government of Karnataka (up to 21 July 2020, after which individual case details were not available in daily bulletins) and linelist contact tracing data maintained by the state’s Integrated Disease Surveillance Programme (up to 1 June 2020). (15) For confirmed COVID-19 cases from 9 March to 21 July 2020 (n=71068), the government bulletins included the unique patient ID, age, sex, district of reporting, case outcome (followed up till 23 August), the unique ID of upstream contact (if known), travel history (if any), and case surveillance definition (if assigned). For confirmed cases from 9 March to 1 June 2020 (n=3404), the IDSP linelist added the symptom status (asymptomatic or symptomatic) at time of sample collection, date of symptom onset (n=261/308 symptomatic cases), date of sample collection and test results, and number of contacts traced (n=956/3404 cases). The two datasets were merged using the unique patient IDs which were common to both datasets. The study followed the Strengthening the Reporting of Observational Studies in Epidemiology (STROBE) reporting guidelines. (16)

### Case identification and contact tracing

The criteria for administering a COVID-19 test were periodically updated by the Indian Council of Medical Research (table S1). (17) A contact was defined as any individual who has been exposed to a confirmed case anytime between 2 days prior to symptom onset (in the positive case) and date of isolation (or maximum 14 days after onset of symptoms). All contacts were quarantined and monitored for 14 days. The duration (>15 minutes), the proximity (<1 meter), and the nature of exposure were taken into consideration when defining a contact. (18) Contacts were further divided into high-risk or low-risk contacts (supplementary S1). A COVID-19 test was administered to all symptomatic contacts and high-risk asymptomatic contacts between day 5 to 10 after contact. (19) For interstate and international travellers, screening was done at all ports of entry including airports, train stations and road checkposts at state borders. Symptomatic travellers were tested and asymptomatic travellers were subject to 14 days of quarantine. (20) (21)

### Case categorisation and ascertainment of transmission pairs

We divided all cases into four mutually exclusive categories based on origin of infection: imported international (history of international travel within 14 days of symptom onset), imported domestic (history of travel from other Indian states into Karnataka), local with known origin (contact with a known COVID-19 case or present at a location or event linked to COVID-19 transmission, and no travel history outside Karnataka), and local cases with unknown origin (no travel history outside Karnataka and could not be traced to any known COVID-19 case or event). Additionally, we categorised cases on the basis of their surveillance case definition at the time of first presentation: Influenza-like illness (ILI: fever ≥38 C° and cough with onset within the last ten days), or Severe acute respiratory infection (SARI: fever ≥ 38C° and cough with onset within the last 10 days and requires hospitalization). This categorisation was independent of the categories based on origin and was assigned for only cases for whom this information was available. If a newly confirmed case had a history of contact with a previously confirmed case, we assumed that the new case acquired the infection from the previously known case; and hence linked them together as secondary case and index case respectively (probable infector-infectee pair). If a secondary case was in contact with 2 or more index cases, we linked the secondary case to the earliest index case.

### Reproduction number (R) and overdispersion parameter (k)

We fit a negative binomial distribution to the observed offspring distribution using a Bayesian Markov Chain Monte Carlo approach and estimated effective reproduction number (R) as the mean, and degree of heterogeneity in transmission as the overdispersion parameter (k) of the fitted distribution. Sample mean and 2.5% and 97.5% percentiles based on post warmup samples were used to evaluate the posterior point estimates and 95% confidence intervals respectively. Since estimates of R and k are sensitive to the completeness of contact tracing, we separately analysed subgroups where contact tracing was known or expected to be comprehensive (supplementary S5 and table S2). Specific metrics of contact tracing adequacy and performance were not available in the dataset. (22) A cluster was defined as two or more confirmed infections with reported close contact. The superspreading threshold was defined as an index case causing eight or more secondary infections. (6,23)

### Secondary attack rate, determinants of risk of infection among contacts and risk of symptomatic infection among cases, and case fatality rate

We computed secondary attack rates by dividing the number of positive contacts by the number of contacts traced. Only high-risk or close contacts were included (supplementary S1). We did a retrospective cohort analysis using a Poisson regression model to estimate adjusted relative risks of infection in contacts, and symptomatic infection in cases as a function of the predictor variables (supplementary S3 and S4). (24) Case fatality rate was calculated as the number of deceased cases divided by total reported cases.

### Serial interval

The serial interval was calculated as the difference between the symptom onset dates of index case and secondary case in each transmission pair. We then fitted parametric distributions (gamma, lognormal, and weibull) to serial interval data using the maximum likelihood method. Akaike Information Criterion (AIC) was used to choose the optimal distribution, and confidence intervals were estimated using 10000 bootstraps.

### Data handling

Data collection was done in Microsoft Office Excel. Data cleaning and analysis was done in Stata 15.0 and Python v3.6.8. Figures were prepared using matplotlib v3.3.2, seaborn 0.10.0, and GraphPad Prism 9. Clusters were visualised using *EpiContacts* package in R. (25)

## Results

From 9 March to 21 July 2020, Karnataka reported 71068 cases with a median age of 37 years (IQR 27-50) of which 63.1% were males. Table 1 and Figure 1 give an epidemiologic summary of the studied outbreak with time and demographic characteristics. Early in the epidemic, most cases occurred in middle-aged adults which can be explained by their high mobility. A ‘V’ shaped pattern can be seen in figure 1C, indicating that more older adults and children were infected with time. The incidence increased throughout the study period, with 55826 (78.55%) cases recorded in July alone. Most cases in March were internationally imported followed by low incidence local transmission in April and May. A majority of the 5105 local cases whose origin was known occurred from April to mid-June. The source of infection of 57670 (81.15%) locally transmitted cases could not be ascertained; most of these cases occurred after mid-June. 20.3% of all cases were detected through symptomatic surveillance (17.1% ILI and 3.2% SARI). The median age of ILI cases was 40 (IQR 29-53) and SARI cases was 54 years (IQR 43-65). The mean delay from symptom onset to lab confirmation (testing delay) was 5.1 (4.7-5.6) days (n=261). Local cases with unknown origin had the longest delay among all categories at 5.7 (5.1-6.2) days (n=112). Detailed results of various delays are given in Table S4.

**Table 1:** Distribution of 71068 COVID-19 cases across time and categories and their demographic characteristics and case fatality rate (Karnataka, India; up to 21 July 2020)

|  | Number of cases, n (%) | Median age (IQR) | Gender Male, % | Case Fatality Rate, % (95% CI) |
| --- | --- | --- | --- | --- |
| **Month** |  |  |  |  |
| March | 101 (0.14%) | 35 (25-50) | 68.32 | 1.98 (0.2 - 7.0) |
| April | 464 (0.65%) | 35 (25-48) | 67.67 | 5.39 (3.5 - 7.8) |
| May | 2656 (3.74%) | 29 (19-39) | 59.94 | 1.13 (0.8 - 1.6) |
| June | 12021 (16.91%) | 34 (24-48) | 62.37 | 3.18 (2.9 - 3.5) |
| July | 55826 (78.55%) | 38 (28-52) | 63.35 | 2.47 (2.3 - 2.6) |
| **Category (origin of infection)** |  |  |  |  |
| Imported Domestic | 7640 (10.75%) | 30 (20-40) | 63.31 | 0.46 (0.3 - 0.6) |
| Imported International | 653 (0.92%) | 35 (25-42) | 76.26 | 0.15 (0.0 - 0.8) |
| Local with known origin | 5105 (7.18%) | 32 (22- 45) | 55.81 | 0.84 (0.6 - 1.1) |
| Local with unknown origin | 57670 (81.15%) | 39 (28-52) | 63.56 | 3.02 (2.9 - 3.2) |
| **Category (surveillance case definition)** |  |  |  |  |
| ILI | 12124 (17.06%) | 40 (29-53) | 68.99 | 2.42 (2.2 - 2.7) |
| SARI | 2295 (3.23%) | 54 (43-65) | 66.54 | 23.66 (21.9 - 25.4) |
| **Total** | **71068** | **37 (27-50)** | **63.09** | **2.54 (2.4 - 2.7)** |

For case fatality rate, month is according to date of case confirmation. ILI=Influenza like illness, SARI=Severe acute respiratory infection.

**Figure 1:**
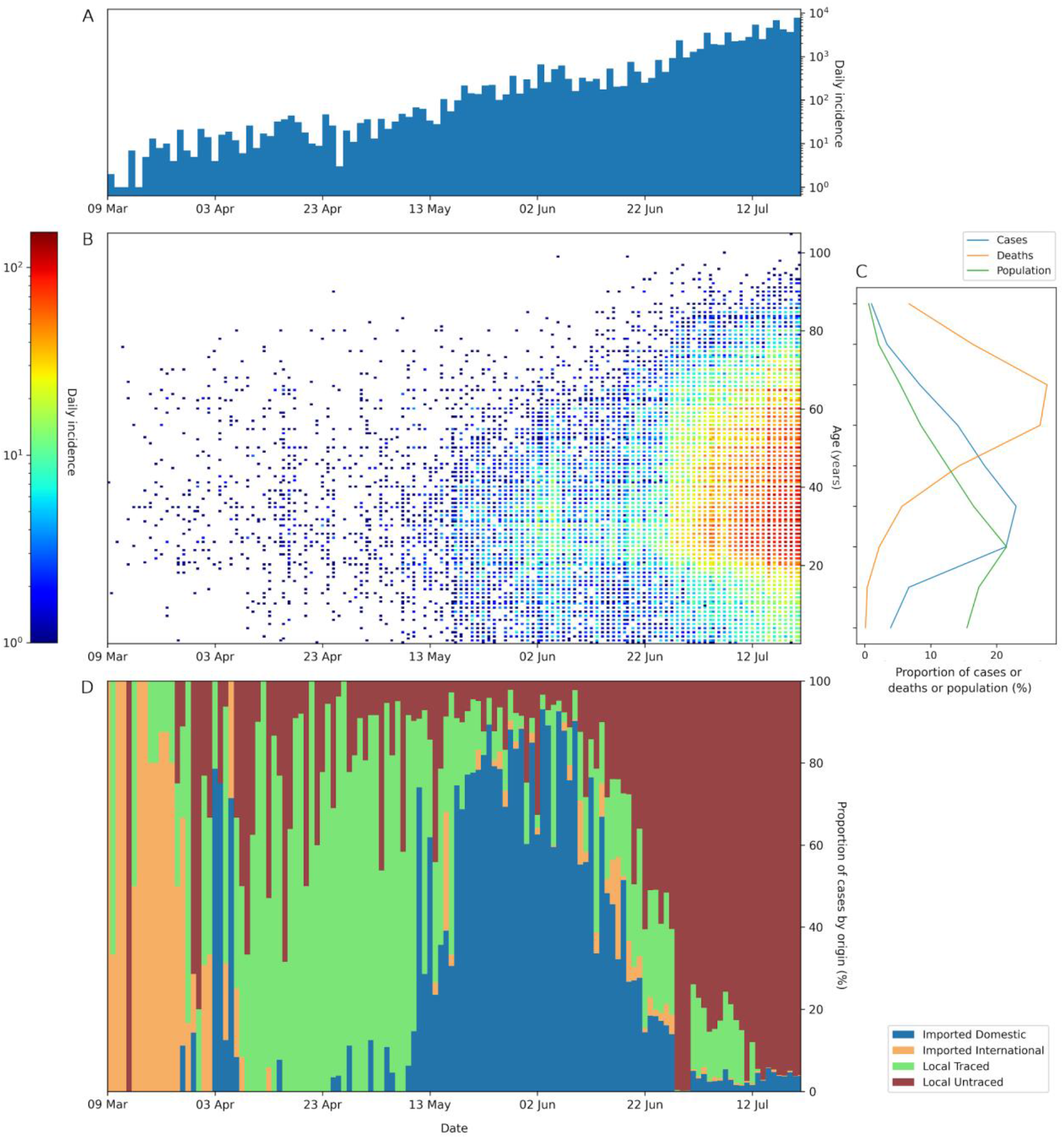
Epidemiological summary of the COVID-19 outbreak in Karnataka, India from 9 March to 21 July 2020 (n=71068). **[A]** Incidence by daily confirmed cases across time. **[B]** Density scatterplot showing daily confirmed cases for various ages across time. **[C]** Distribution of population, COVID-19 cases and COVID-19 deaths across age groups. 10-year age brackets have been used starting from 0-9 years, and the last bracket is ≥80 years. **[D]** Daily confirmed cases across time stratified by origin of infection-imported international, imported domestic, local with known origin, and local with unknown origin.

### Overdispersion (k), reproduction number (R), and major transmission clusters

We found 111 instances where the index case crossed the super-spreading threshold of ≥8 secondary cases upto 21 July 2020 in Karnataka. A total of 1277 clusters were identified with 6424 linked cases till 21 July 2020 in Karnataka. There were 7 clusters with a size larger than 50 cases and 106 clusters with more than 10 cases each. We characterised the three largest clusters including the Bellary cluster (221 cases), the Delhi convention cluster (97 cases), and the Pharmaceutical company cluster (76 cases). The reconstructed transmission networks for these three clusters are shown in Figure 2. For 394 cases linked to these three largest clusters, the estimated R was 0.91 (0.72-1.15) and k was 0.22 (0.17-0.27). 12.4% of infectious cases caused 80% of all transmission, while 71.2% of cases did not lead to any further transmission among these three clusters (figure 3D). In late-March, a cluster of infections was seeded by travellers returning from the religious convention at Nizammudin mosque in Delhi. In the Bellary cluster, the infection was seeded by a worker at a steel plant in Bellary (26), followed by rapid spread among the employees living in close-contact in dormitories. The initial infections occurred in the management department which had a high contact rate with employees from other departments thus facilitating spread. A similar cluster was reported in Nanjangud, Mysore (27) where a pharmaceutical company became the origin of a large cluster.

**Figure 2:**
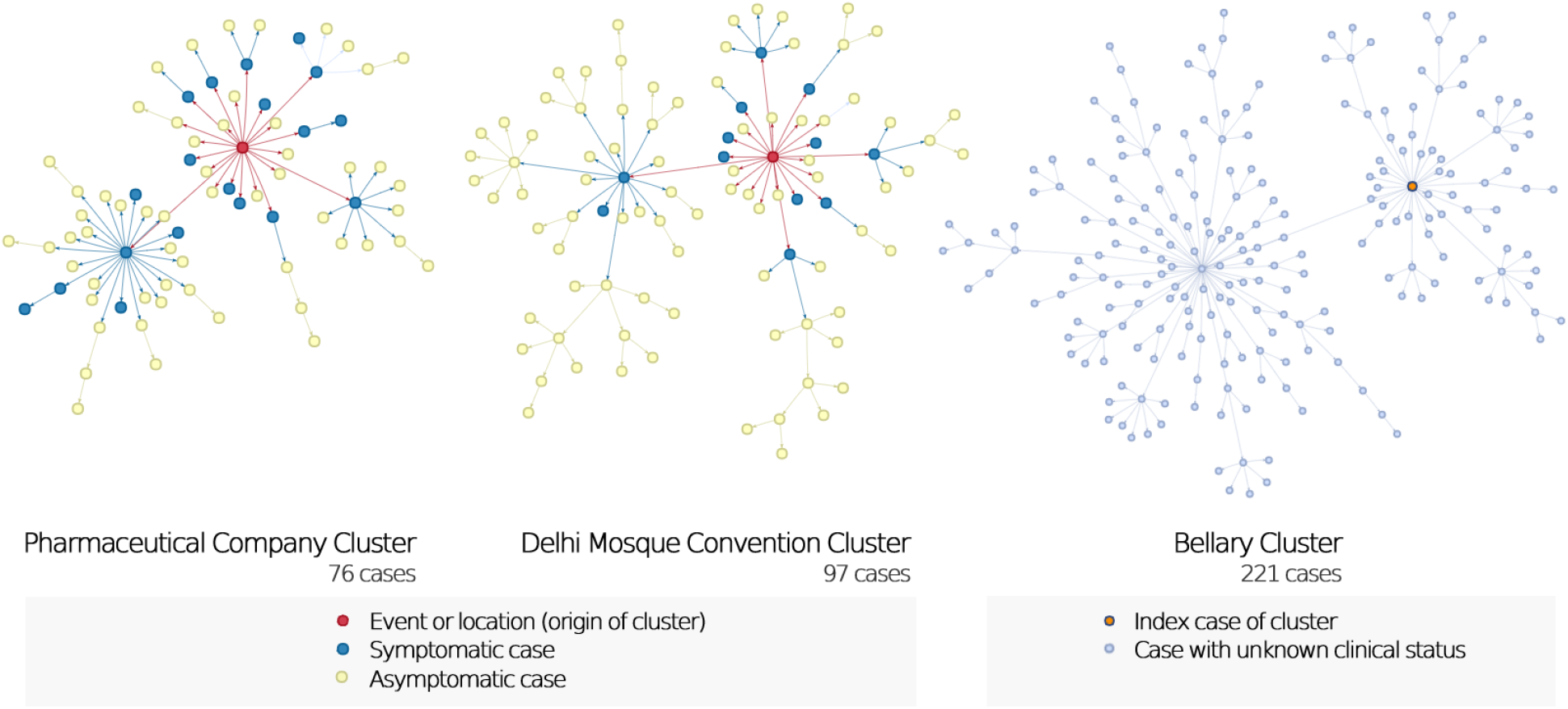
Transmission networks of three largest clusters of SARS-CoV-2 cases in Karnataka, India up to 21 July 2020. The networks indicate the heterogeneity in transmission from infected cases, with a few patients causing most secondary cases. The Bellary cluster occurred in June and July, during which symptomatic status of cases was not available.

**Figure 3:**
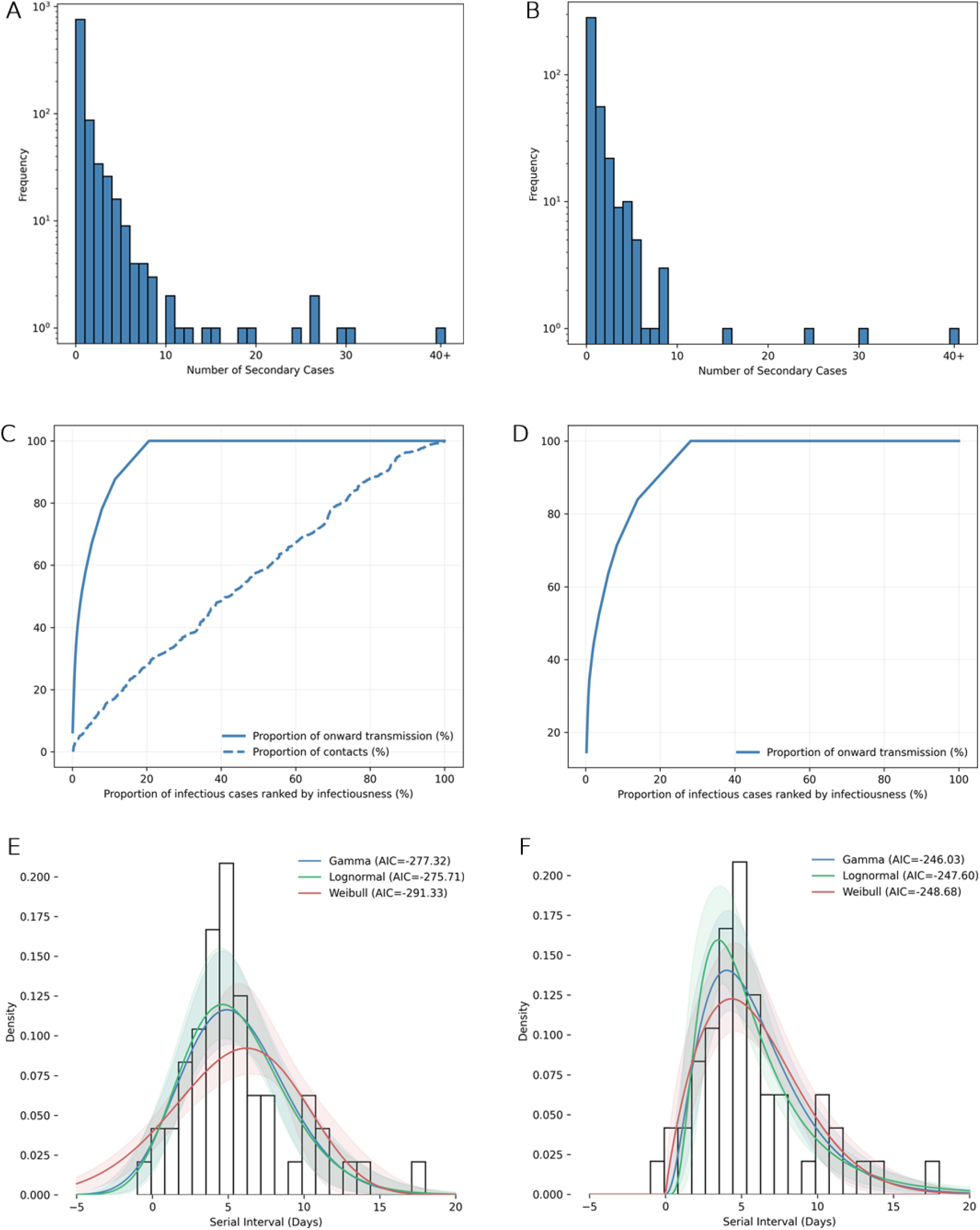
Transmission characteristics of SARS-CoV-2 in Karnataka, India. **[A**,**B]** Observed offspring distribution of [A] 956 cases with confirmed forward contact tracing and [B] 394 cases linked to the three largest clusters (Bellary cluster, Delhi convention cluster, and Pharmaceutical company cluster). Bars show observed frequency of the number of individuals infected by each case. **[C**,**D]** Proportion of all onward transmission (solid line) and proportion of all contacts (dashed line) due to a given proportion of infectious cases, where cases are ranked by infectiousness, for [C] 956 cases with confirmed forward contact tracing and [D] 394 cases linked to the three largest clusters. **[E**,**F]** Serial interval distribution of SARS-CoV-2 infections in Karnataka, India and the fitted gamma, lognormal and weibull distributions, among [E] 53 infector-infectee pairs and [F] 51 infector-infectee pairs with positive serial intervals. 10000 bootstraps were used for estimating the 95% confidence limits shown by the shaded bands.

For 956 cases whose contacts were confirmed to have been traced, the estimated R was 0.75 (0.62-0.91) and k was 0.12 (0.11-0.15). 8.7% of infectious cases had 14.4% of contacts but caused 80% of all transmission (figure 3C). On the other hand, 79.4% of cases did not lead to any further transmission despite having 71.6% of all recorded contacts. Asymptomatic cases had a much lower R than symptomatic cases (0.41 [0.32-0.52] vs 2.04 [1.56-2.67]), but had higher transmission heterogeneity indicated by a lower k (0.12 [0.09-0.15] vs 0.29 [0.23-0.37]). For all cases till 13 June (n=6824), R was 0.23 (0.20-0.26) and k was 0.04 (0.03-0.04). Complete results for R and k are given in supplementary S5 and table S2.

### Secondary attack rate and determinants of risk of infection among contacts

The secondary attack rate (SAR) among all close contacts was 3.6% (95%CI, 3.4-3.9). Results are summarised in table 2 and 3. The median number of contacts identified for an index case were 11 (IQR 5-21). The median number of contacts per case were higher (15 [IQR 8-35]) when the index case was confirmed ≥4 days after symptom onset. Symptomatic index cases were more infectious than asymptomatic cases (SAR 7.7% vs 2.0%; aRR 3.63 [3.04-4.34]). As compared to infectors aged 19-44 years, children were less infectious even after controlling for other factors including symptomatic status (aRR=0.21 [0.07-0.66] for 0-5 years and 0.47 [0.32-0.68] for 6-18 years). Adults aged ≥45 years seemed to be more infectious, but this association disappeared when controlling for increased symptomaticity of older adults. Male infectors were found to be less infectious than females (aRR 0.78 [0.66-0.92]). Infectors who had a delay from symptom onset to confirmation of ≥4 days were associated with higher infectiousness (aRR=3.01 [2.11-4.31]). Local cases with unknown origin had the highest infectivity of the four categories by origin, with R of 1.04 (0.87-1.25) and SAR of 8.5% (7.7 - 9.4). Cases identified through symptomatic surveillance had the highest SAR of any subgroup (14% and 9.8% for ILI and SARI respectively).

**Table 2:** Secondary attack rate of SARS-CoV-2 among 16715 contacts of 956 index cases (Karnataka, India; up to 1 June 2020)

|  | Median number of contacts<br>per index case, n (IQR) | Positive contacts/<br>Total contacts | SAR, % (95%CI) |
| --- | --- | --- | --- |
| **Month** |  |  |  |
| March | 10.5 (6-21.25) | 59/1408 | 4.2 (3.2 - 5.4) |
| April | 12 (6-27) | 349/7120 | 4.9 (4.4 - 5.4) |
| May | 10 (5-28) | 202/8187 | 2.5 (2.1 - 2.8) |
| **Symptom status of index case** |  |  |  |
| Symptomatic | 13 (7-28) | 367/4788 | 7.7 (6.9 - 8.4) |
| Asymptomatic | 10 (5-20) | 243/11927 | 2.0 (1.8 - 2.3) |
| **Category (origin of infection) of index case** |  |  |  |
| Imported international | 10 (5.25-18.5) | 24/1075 | 2.2 (1.4 - 3.3) |
| Imported domestic | 9 (3-20) | 47/4670 | 1.0 (0.7 - 1.3) |
| Local with known origin | 11 (6-20) | 178/6733 | 2.6 (2.3 - 3.0) |
| Local with unknown origin | 12 (7-13) | 361/4237 | 8.5 (7.7 - 9.4) |
| **Category (surveillance case definition) of index case** |  |  |  |
| ILI (not requiring hospitalisation) | 17 (9-31) | 87/622 | 14.0 (11.4 - 17.0) |
| SARI (requiring hospitalisation) | 16 (10-43) | 134/1371 | 9.8 (8.3 - 11.5) |
| **Delay from symptom onset to confirmation of index case** |  |  |  |
| <4 days | 10.5 (6-20.25) | 41/1091 | 3.8 (2.7 - 5.1) |
| ≥4 days | 15 (8-35) | 313/3256 | 9.6 (8.6 - 10.7) |
| **Overall** | **11 (5-21)** | **610/16715** | **3.6 (3.4 - 3.9)** |

**Table 3:** Risk of SARS-CoV-2 infection among 16715 contacts of 956 index cases (Karnataka, India; up to 1 June 2020)

|  | Crude RR (95%CI) | Adjusted RR* (95%CI) |
| --- | --- | --- |
| **Age group of index case** |  |  |
| 0-5 years | 0.22 (0.07- 0.69) | 0.21 (0.07 - 0.66) |
| 6-18 years | 0.40 (0.27 - 0.59) | 0.47 (0.32 - 0.68) |
| 19-44 years | 1 (reference) | 1 (reference) |
| 45-64 years | 1.47 (1.22- 1.77) | 1.02 (0.85- 1.23) |
| ≥65 years | 1.45 (1.16- 1.82) | 0.89 (0.70- 1.13) |
| **Sex of index case** |  |  |
| Female | 1 (reference) | 1 (reference) |
| Male | 0.89 (0.76 -1.05) | 0.78 (0.66 - 0.92) |
| **Symptom status of index case** |  |  |
| Asymptomatic | 1 (reference) | 1 (reference) |
| Symptomatic | 3.76 (3.21- 4.41) | 3.63 (3.04- 4.34) |
| **Delay from symptom onset to lab confirmation in index case \*\*** |  |  |
| <4 days | 1 (reference) | 1 (reference) |
| ≥4 days | 2.56 (1.86- 3.52) | 3.01 (2.11- 4.31) |

\* Adjusted for age group, sex and symptom status of index case.
\*\* Adjusted for age group and sex of symptomatic index case. This predictor variable was part of a separate model that included only symptomatic index cases and their contacts.

### Serial Interval

After excluding asymptomatic cases, 54 infector-infectee pairs were identified where symptom onset dates for both infector and infectee were available. 35, 14 and 5 pairs were from March, April and May respectively. One pair was dropped for having a large negative serial interval (−19 days). Estimated parameters for the serial interval distribution are shown in table S3 and the fit is shown in figures 3E,F and S3. When analysing the 53 pairs, Weibull distribution was the best fit (AIC=-291) with a mean of 5.4 (4.4-6.4) and SD of 4.3 (3.0-5.1) days. When analysing serial intervals with positive values only (51 pairs), the Weibull distribution was again the best fit (AIC=-249) with a mean of 5.9 (5.0-6.9) and SD of 3.3 (2.4-4.1) days.

### Determinants of presence of clinical symptoms in SARS-CoV-2 infection

Among 3404 cases till 1 June, 9.0% were symptomatic at the time of sample collection. Increasing age, male sex, and having a symptomatic infector were associated with a higher probability of being symptomatic given SARS-CoV-2 infection (Table 4). The probability of symptomatic infection increased with age. When compared to cases aged 19-44 years old, children were about half as likely to be symptomatic (aRR=0.46 [0.19-1.11] for 0-5 years and 0.39 [0.24-0.64] for 6-18 years); whereas older adults aged 45-64 years were about twice as likely to be symptomatic (aRR=2.24 [1.77-2.84]), and elderly aged ≥65 years were about four times as likely to be symptomatic (aRR=4.46 [3.37-5.90]). Males were 1.29 (1.04-1.62) times more likely to be symptomatic than females. A secondary case was 8.16 (3.29-20.24) times more likely to be symptomatic if the index case was also symptomatic compared to if it was asymptomatic. Presence of one or more comorbidities was associated with increased symptomaticity but was not statistically significant (aRR=1.13 [0.92-1.39]).

**Table 4:** Symptomatic proportion and risk of developing symptoms among 3404 COVID-19 cases (Karnataka, India; up to 1 June 2020)

|  | Symptomatic cases, n (%) | Crude RR (95%CI) | Adjusted RR* (95%CI) |
| --- | --- | --- | --- |
| **Age group of case** |  |  |  |
| 0-5 years (n - 147) | 5 (3.4) | 0.46 (0.19 - 1.09) | 0.46 (0.19 - 1.11) |
| 6-18 years (n - 602) | 17 (2.8) | 0.38 (0.23 - 0.62) | 0.39 (0.24 - 0.64) |
| 19-44 years (n - 1955) | 146 (7.4) | 1 (reference) | 1 (reference) |
| 45-64 years (n - 570) | 96 (16.8) | 2.26 (1.77- 2.87) | 2.24 (1.77- 2.84) |
| $\geq 65$ years (n - 130) | 44 (33.8) | 4.53 (3.40 - 6.03) | 4.46 (3.37- 5.90) |
| **Sex of case** |  |  |  |
| Female (n - 1309) | 97 (7.4) | 1 (reference) | 1 (reference) |
| Male (n - 2095) | 211 (10.1) | 1.36 (1.08 - 1.71) | 1.29 (1.04- 1.62) |
| **Comorbidity in case** |  |  |  |
| Present (n - 1043) | 117 (11.2) | 1.39 (1.11- 1.72) | 1.13 (0.92- 1.39) |
| Absent (n - 2361) | 191 (8.1) | 1 (reference) | 1 (reference) |
| **Symptom status of index case\*\*** |  |  |  |
| Asymptomatic (n - 370) | 5 (2.9) | 1 (reference) | 1 (reference) |
| Symptomatic (n - 455) | 55 (12.1) | 8.95 (3.62- 22.11) | 8.16 (3.29 - 20.24) |
| **Overall (n - 3404)** | **307 (9.0)** | **-** | **-** |

\*Adjusted for age group, sex and presence of comorbidity in affected case, and symptom status of the index case.
\*\*Symptom status of index case was not available for 2579 out of 3404 cases whose infector was unknown (these cases were included in the model by making a separate category not shown in the table).

### Case Fatality Rate

The case fatality rate (CFR) across time and case categories is presented in Table 1 and across age and sex in Figure 4. Among 71068 cases, the overall CFR was 2.5% (95% CI, 2.4-2.7%). Local cases with unknown origin had the highest CFR (3.0%), followed by local cases with known origin (0.84%), imported domestic cases (0.46%), and imported international cases (0.15%). In surveillance case categories, SARI cases had a much higher CFR (23.66%) than ILI (2.42%). CFR steadily increased with age in both sexes, ranging from 0.07% in 0-9 years old to 16.31% in males and 17.55% in females of age 80 and above (Figure 4). The highest CFR was recorded for cases that were reported in the month of April (5.39%).

**Figure 4:**
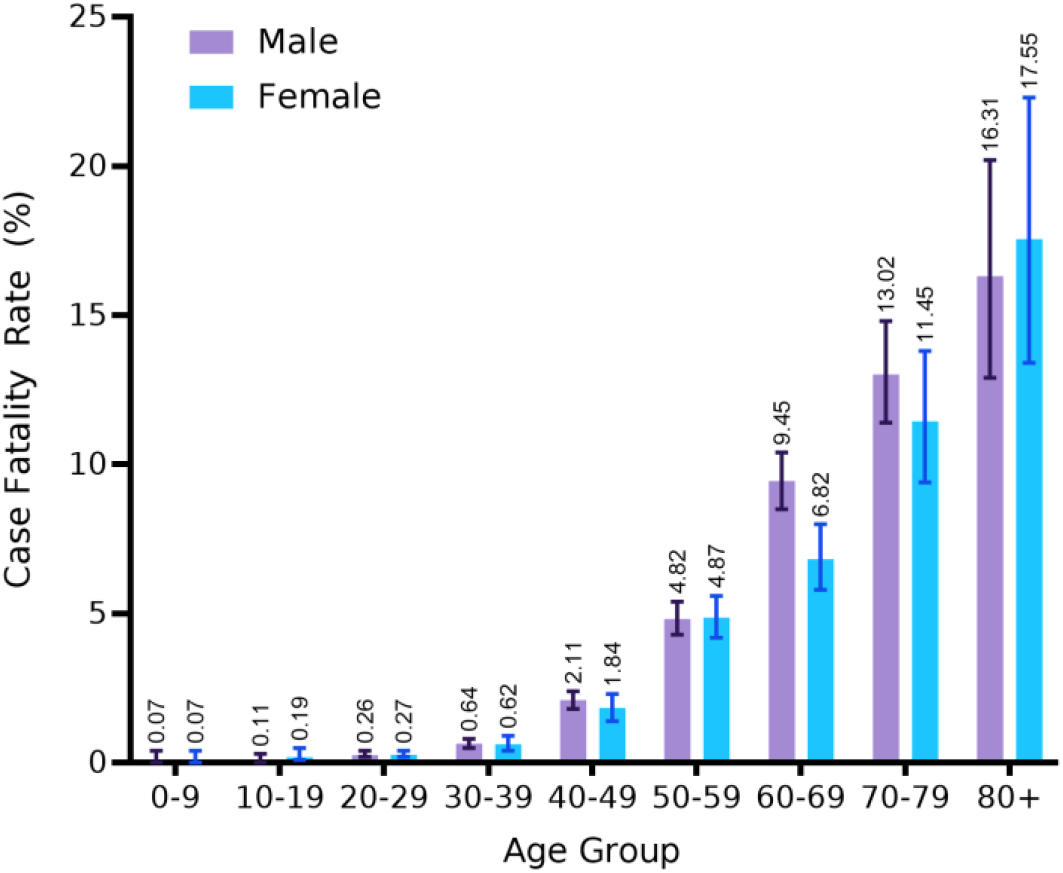
Case fatality rate COVID-19 cases stratified by age and sex in Karnataka, India up to 21 July 2020 (n-71068). Bars represent mean CFR for that subgroup (value shown above bars) with error bars showing the 95% confidence intervals.

## Discussion

Although the reproduction number R0 of the novel coronavirus has been well characterised, we find that R0 alone fails to capture the true picture of individual-level transmission dynamics. Overdispersion (k) ranged from 0.04 to 0.34 in our study, confirming that there is significant heterogeneity in transmission of the novel coronavirus. Importantly, we did not find significant underlying heterogeneity in the number of contacts. Figure 3C shows clustering in the number of secondary cases (concave line) while the number of contacts are homogenous as indicated by a relatively linear relation. Heterogeneity in transmission can be explained by heterogeneity in the number of contacts and/or the probability of infection per contact (infectivity level of index case and nature of exposure).(28,29) Modelling studies indicate that SARS-CoV-2 SSEs occur when an infected person is briefly shedding at a very high viral load and has a high concurrent number of exposed contacts. (30) Since the patients which caused a majority of secondary cases did not have a concurrent larger share of total contacts (8.7% of infectious cases had 14.4% of contacts but caused 80% of transmission), our findings underscore the importance of the high infectivity of index case (at the time of exposure) and the nature of exposure in causing a successful SSE.

The most reliable estimates of overdispersion (k) in our study were 0.12 (0.11-0.15) for confirmed traced cases (n=956) and 0.22 (0.17-0.27) from the three clusters (n=394). Our estimates of k align with the lower range of current global estimates, albeit with smaller confidence intervals due to larger sample size. A modelling study analysing global clusters estimated k at 0.10 (0.05-0.20). (31) A study of 1288 cases estimated k at 0.06 (0.05-0.07) and 0.20 (0.09-0.31) in two states of Indonesia. (32) In China, k was estimated at 0.25 (0.13–0.88) from 135 cases in Tianjin (33) and at 0·58 (0·35–1·18) from 391 cases in Shenzhen. (34) In Hong Kong, k was estimated at 0.43 (0.29–0.67) from 290 cases. (23) In Georgia USA, the overall k ranged from 0.32 to 0.49, with even lower values after shelter-in-place orders were issued. (35) Our findings suggest that super-spreading played a more dominant role in transmission in Karnataka, India, as compared to most high-income countries. Interestingly, phylogenetic studies indicate that remote clusters can be retrospectively linked to a previous SSE, indicating that super-spreading may play an even larger role in the overall propagation of the epidemic than is detected through surveillance and tracing. (36) Ideally then, the true picture of epidemics can only be understood by analysing contact tracing, serological and phylogenetic data together, which can help plan adequate control measures.

Modelling studies indicate that the delay from symptom onset to confirmation (testing delay) is an essential determinant of the effectiveness of contact tracing. (5,37) Indeed, we found that infectors diagnosed four or more days after symptom onset led to a higher SAR among their contacts (9.6% vs 3.8%) and also had a higher number of contacts (median 15 vs 10.5). We found that this delay was lower for cases who had an early first contact with surveillance or contact tracing systems, namely, cases screened at entry ports (imported cases) and local cases from known contact lists. Imported cases had a low reproduction number compared to local cases (Table S2) which suggests that screening and registration of all persons at entry ports leading to early detection of these cases prevented most onward transmission. Cases captured through symptom-based surveillance and those whose source of infection was unknown had a higher SAR than all other case categories (Table 2) and had a concurrently higher testing delay (table S4). SARI had lower infectiousness than ILI cases which could be explained by their hospitalisation preventing some transmission. Cases that were from known contact lists were confirmed 0.88 days (0.60-2.04) earlier and caused 60% less secondary infections than local cases whose origin was unknown (R 1.04 vs 0.42). These findings highlight the effect of contact tracing in reducing transmission and reassert the importance of minimising testing and tracing delays.

We estimated the relative risk (aRR) of infectivity at 3.63 (3.04-4.34) for symptomatic infectors as compared to asymptomatic carriers (SAR 7.7 vs 2.0%; R 2.04 vs 0.41) (table 2 and S2). Other studies have found that infectiousness also increases with disease severity. (38) This suggests that predictors of symptomatic infection and/or severe disease (like age) are by extension also predictors of increased infectiousness (Table 4). Accordingly, interventions aimed at children might have a relatively small impact on reducing SARS-CoV-2 transmission, as suggested by their low symptomaticity and severity and thus lower infectiousness than adults. (39) Our study also finds that children aged 6-18 are less infectious even after adjusting for lower symptomatic infections in this group. Interestingly, we found a strong association between the symptom status of the index and secondary case; symptomatic infectors were 8.16 (3.29-20.24) times more likely to generate symptomatic secondaries. These findings are corroborated by He D et al., who estimated relative risk of infectivity of symptomatics against that of asymptomatics at 3.9 (1.5–11.8), and at 6.6 (2.0–34.7) when focusing on symptomatic secondaries. (40) With symptomatic infectors both, more likely to produce secondary cases in general and also more likely to produce symptomatic secondaries who are themselves more infectious, it seems that a cascading effect of high transmission potential may play a role in amplifying COVID-19 outbreaks. Though it is known that nasopharyngeal viral load is higher in symptomatic cases and increases with severity in COVID-19, (41) it would be pertinent to explore whether a higher infectious dose from the infector influences symptom status and infection severity in the secondary case, something that could explain findings from this and He D et al. (40,42)

We found one study estimating the serial interval of SARS-CoV-2 from contact tracing data in India; however, this study assumed the date of sample collection in asymptomatic cases as a proxy for symptom onset which heavily affects the reliability of their estimate. (43) Although we present reliable estimates of serial interval for COVID-19 for the first time from India, enhanced data sharing enabling real-time estimation to inform policy decisions is recommended to account for the temporal variation of serial interval as observed in China. (44) Given that the reproduction number is sensitive to the value of serial interval used for its estimation, it is prudent to select the serial interval distribution that fits best in context of the location and time phase of the epidemic. (45) Our estimated mean serial interval of 5.4 (4.4-6.4) agrees with existing evidence from global studies. (10)

Our study has certain limitations. Firstly, symptomatic status was based on data collected at the time of sample collection and hence some cases recorded as asymptomatic may have developed symptoms later. (46–48) This would overestimate the proportion of asymptomatic infections and also the relative transmissibility by asymptomatics since presymptomatic cases have been shown to be more infectious than asymptomatic carriers. (40,47,49) Second, any amount of case and/or contact under-ascertainment during surveillance and contact tracing carries the potential to bias our results. Although we have attempted to minimise this bias by analysing subgroups with high reliability of data (Table S2), some degree of bias can still be expected. Since the completeness of contact tracing is inherently limited by memory recall and logistics, R estimated from contact tracing data cannot capture the entirety of the outbreak and can thus be expected to underestimate the true R. Third, details of settings of transmission and timing of exposure of contact to index case were not available for the vast majority of cases which precluded any insightful analysis on the same. Finally, the dates of symptom onset in our study may be subject to recall bias. Additionally, our results should be interpreted in the context of the NPIs in place at the time of the study. From 25 March to 8 June, physical distancing was mandated in Karnataka with closures of schools and most public places, restrictions on large gatherings, and mask wearing in public settings.

Our findings have a few important implications for optimizing policy. We find that even though surveillance, tracing, and social distancing may keep the reproduction number and hence transmission at low levels, super spreading is common in COVID-19 and carries the potential to acutely overwhelm surveillance and tracing systems. However, this presents an opportunity as well, in that outbreaks where a minority of cases cause most further transmission (high dispersion, low k) are much more amenable to control through measures that target the high-risk groups and settings responsible for most of the transmission. (6–8) Existing measures that limit potential super-spreading including bans on large gatherings and capping capacity in closed spaces are expected to remain beneficial. (28) Specifically, emphasis should be given on backward or retrospective contact tracing which becomes increasingly effective as overdispersion increases, and tracing and testing delays should be minimized. (31,37) Evidence on the effect of symptom status on transmission suggests that measures targeted at children will not reduce transmission significantly. (39,49) Although more studies are needed, there is increasing evidence that symptomatic cases beget more symptomatic secondaries and may cause a snowballing effect on transmission across generations, (40) which has significant implications for both-transmission and morbidity control in COVID-19 outbreaks.

## Data Availability

The bulletins dataset and all codes used to produce the results are available at www.github.com/CovidToday/covid19-karnataka . Interactive visualisations of cluster transmission networks can be found at www.kaclusters.covidtoday.in . The IDSP dataset can be made available upon reasonable request to the office of Commissioner (PP).

https://kaclusters.covidtoday.in

## Contributors

MG conceptualized the study. MG and GGP supervised and coordinated the study. JA and PP oversaw and monitored the contact tracing activity and data collection in Karnataka and finalized the daily bulletin that was mined for the analysis. GRB and VR acquired contact tracing data from the health department. MSS, GGP, BB, MG, NA cleaned and created the composite dataset. MG, RM, GGP, MSS did the formal analyses. DP, RM, MG, BB, MSS made the figures. GGP, MG, MSS, BB, MA, NA, AM, AG reviewed relevant scientific literature. GRB, VR and TB consulted on the analyses. PP and JA verified the underlying data. MG, AG, GGP, AM, RM, MA, and MSS prepared the manuscript draft. All authors interpreted the results, contributed in revising the draft for important intellectual content and approved the final version for submission.

## Acknowledgement

We thank the India COVID-19 Apex Research Team (iCART) for enabling this research. GRB is funded by an Intermediate Fellowship by the Wellcome Trust DBT India Alliance (Clinical and Public Health research fellowship); grant number IA/CPHI/14/1/501499. We also thank one of the developers of www.covidtoday.in, Mr. Pratik Mandlecha for sharing his expertise. We thank the multi-sectoral contact tracing and surveillance teams and the Health Department of Karnataka whose efforts made this study possible.

## Ethics approval

Surveillance and contact tracing activities wherein the data was collected were part of ongoing outbreak investigation mandated by state and national health authorities. The requirement for full review was waived by the Institutional Ethics Committee, Indian Institute of Public Health, Bengaluru of Public Health Foundation of India (IIPHHB/TRCIEC/211/2020 dated 25/12/2020).

## Data sharing and code availability

The bulletins dataset and all codes used to produce the results are available at github.com/CovidToday/covid19-karnataka. Interactive visualisations of cluster transmission networks can be found at kaclusters.covidtoday.in. The IDSP dataset can be made available upon reasonable request to the office of Commissioner (PP).

## Supplementary Materials

### S1. Definition of high risk and low risk contact ^1^

#### High-risk contact

- Lives in the same household as the case
- Anyone in close proximity (within 1 meter) of the confirmed case without precautions
- Touched or cleaned the linens, clothes, or dishes of the patient.
- Had direct physical contact with the body of the patient including physical examination without PPE.
- Passenger in close proximity (within 1 meter) of a conveyance with a symptomatic person who later tested positive for COVID-19.
- Touched body fluids of the case without appropriate PPE (respiratory tract secretions, blood, vomit, saliva, urine, feces)

#### Low-risk contact

- Any contact not fitting into the above high-risk contact description.

### S2. Bayesian Markov Chain Monte Carlo sampling for estimating reproduction number and overdispersion from contact tracing data

We implemented a Bayesian Markov Chain Monte Carlo sampling method using the NUTS sampler in PyStan 2.18.0.0 with gamma prior estimates of R and k with mean 2.5 SD 2.0, and mean 0.45 SD 0.1 respectively based on previous studies. ^2–5^ 8 chains were parallelly simulated for 5000 iterations and were checked for convergence of Rhat to 1. The first 500 iterations for all chains were discarded as a burn-in period.

### S3. Determinants of risk of infection among contacts

We fit a Poisson regression model with robust error variance to data on contacts’ recorded infection status (binary outcome: contact infected OR not infected) and attributes of each contact’s index cases to estimate adjusted relative risks for infection of contacts as a function of these exposures.^6^ For a subset of this data that included only symptomatic index cases and their contacts, we fit another model which also included the delay from symptom onset to confirmation of the index case (a proxy for delay in case isolation and increasing infectious period spent in community) as a predictor variable. Only high-risk/close contacts were included because the recording of ‘low-risk contact’ status was not deemed reliable in the accessed dataset.

### S4. Determinants of presence of clinical symptoms in SARS-CoV-2 infection

We fit a Poisson regression model with robust error variance to data on confirmed COVID-19 cases’ symptom status (binary outcome: case is symptomatic OR asymptomatic) and attributes of each case and their index case (where known) to estimate adjusted relative risks for developing symptoms in a COVID-19 case as a function of these exposures.^6^

### S5. Minimizing bias due to incomplete contact tracing

The accuracy of reproduction number (R) and overdispersion (k) estimates from data depends on the completeness and adequacy of the contact tracing efforts during the sampled time. For example, cases with zero linked secondary cases in the dataset could either be due to no further transmission from the case after adequate tracing, or due to inadequate or absent contact tracing of the case leading to failure of identification of secondary cases. To account for the possible bias arising out of inadequate contact tracing specially at later stages of the study, we estimated R and k for multiple time periods, and for cases from each category and from major clusters separately.

Three cut-off dates are evaluated in table S2 and figure S2, with the following rationale.

- 1 June was the last date for which the detailed IDSP dataset was available (refer to “Data sources” under Methods section).
- As the epidemic progressed, contact tracing systems were stretched leading to inadequate identification or follow-up of contacts of some cases. We assumed that prevailing contact tracing was adequate if the origin of at least 80% of daily cases was known.^7,8^ This threshold was breached consistently (for more than seven days in a row) after 13 June. This became our second cut-off date for R and k analysis (to minimize bias due to inadequate tracing after this date).
- 21 July was the last date for which the state government bulletins were available with individual case details (refer to “Data sources” under Methods section).

### S6. Additional details on surveillance and tracing methods utilized by the state of Karnataka, India

Karnataka utilised a technology backed multi-sectoral approach to contact tracing with collaboration between health department and law enforcement personnel to effectively trace and track cases. Innovations in quarantine enforcement included a Quarantine Watch mobile app and community based Mobile Squads to monitor contacts in quarantine. It was made mandatory for all returnees and travellers coming to Karnataka to register on a web portal, which enabled comprehensive screening at all entry points and adequate follow up after entry. Additionally, the state carried out a physical and phone-based survey which covered 15M out of 17M households in Karnataka to identify and protect high-risk populations including the elderly, pregnant females, persons with comorbidities, and persons with an ILI or SARI case profile.

**Table S1.**
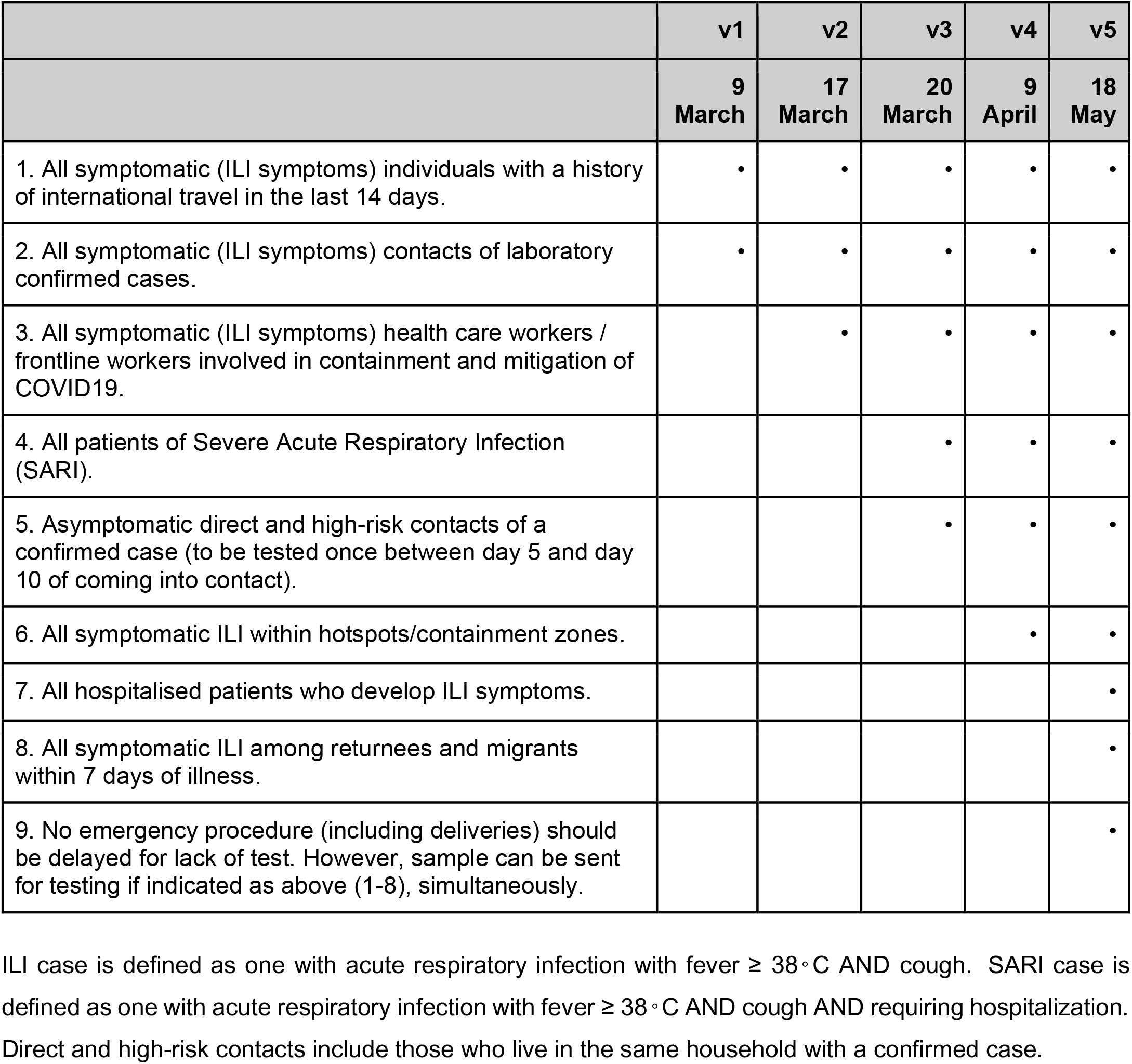
Criteria for testing of COVID-19 in various versions of the recommended strategy by the Indian Council of Medical Research (ICMR) ^9^.

|  | v1 | v2 | v3 | v4 | v5 |
| --- | --- | --- | --- | --- | --- |
|  | 9<br>March | 17<br>March | 20<br>March | 9<br>April | 18<br>May |
| 1. All symptomatic (ILI symptoms) individuals with a history of international travel in the last 14 days. | • | • | • | • | • |
| 2. All symptomatic (ILI symptoms) contacts of laboratory confirmed cases. | • | • | • | • | • |
| 3. All symptomatic (ILI symptoms) health care workers / frontline workers involved in containment and mitigation of COVID19. |  | • | • | • | • |
| 4. All patients of Severe Acute Respiratory Infection (SARI). |  |  | • | • | • |
| 5. Asymptomatic direct and high-risk contacts of a confirmed case (to be tested once between day 5 and day 10 of coming into contact). |  |  | • | • | • |
| 6. All symptomatic ILI within hotspots/containment zones. |  |  |  | • | • |
| 7. All hospitalised patients who develop ILI symptoms. |  |  |  |  | • |
| 8. All symptomatic ILI among returnees and migrants within 7 days of illness. |  |  |  |  | • |
| 9. No emergency procedure (including deliveries) should be delayed for lack of test. However, sample can be sent for testing if indicated as above (1-8), simultaneously. |  |  |  |  | • |

ILI case is defined as one with acute respiratory infection with fever $\geq 38^{\circ}\text{C}$ AND cough. SARI case is defined as one with acute respiratory infection with fever $\geq 38^{\circ}\text{C}$ AND cough AND requiring hospitalization. Direct and high-risk contacts include those who live in the same household with a confirmed case.

**Table S2.**
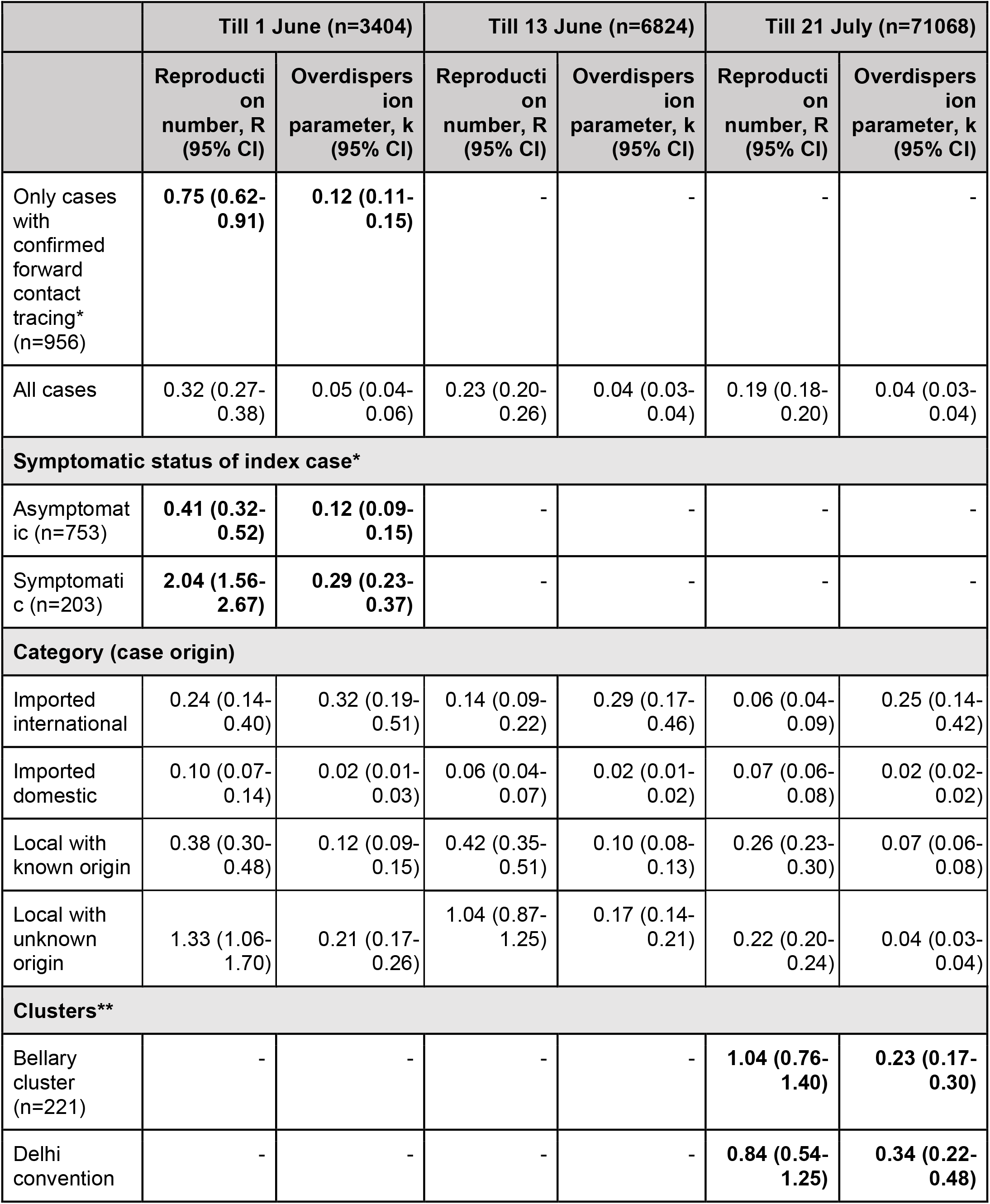
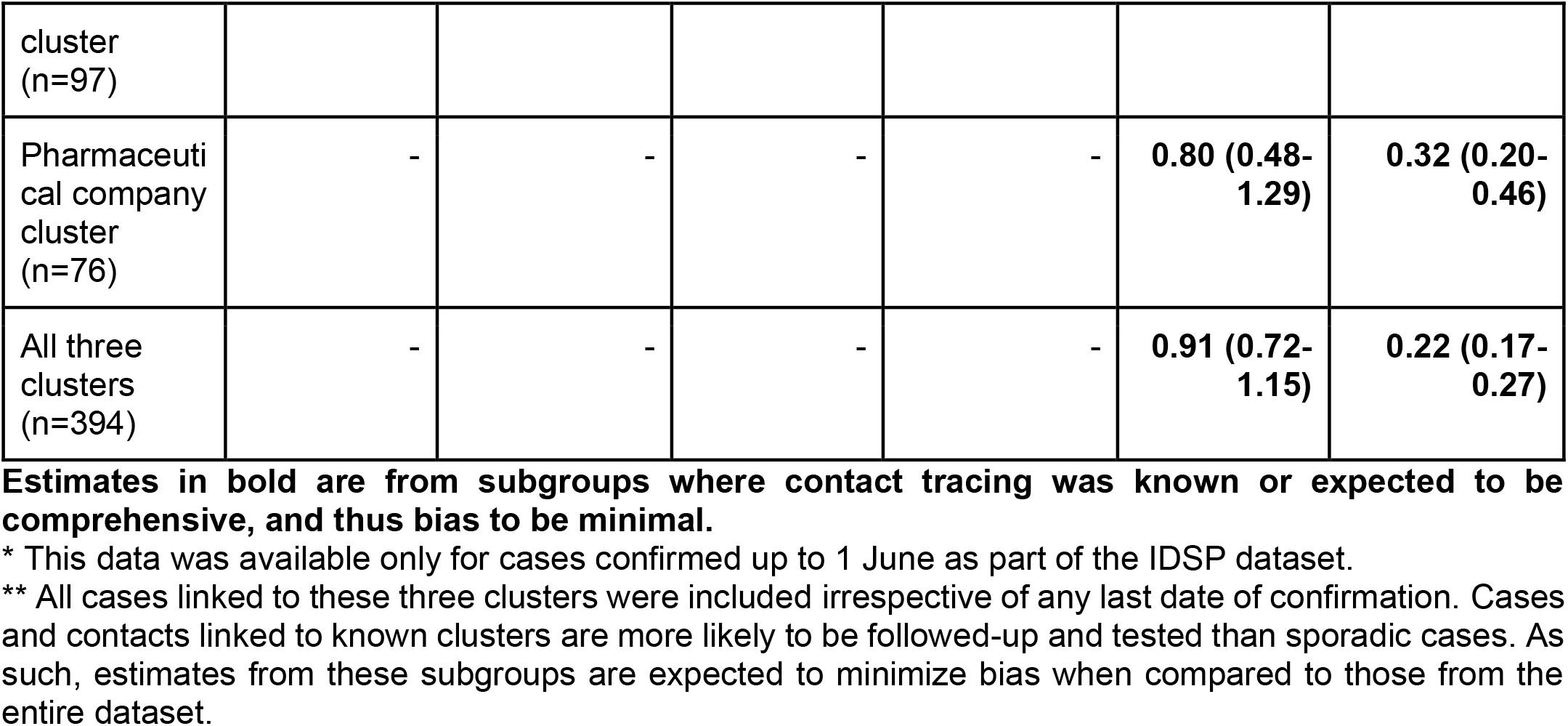
Posterior estimates of reproduction number (R) and overdispersion parameter (k)

**Table S3:** Serial interval of SARS-CoV-2 estimated by fitting parametric distributions to data.

| Distribution | Mean | Standard Deviation | Median | IQR | Goodness of fit (AIC)* |
| --- | --- | --- | --- | --- | --- |
| **All values of serial interval (53 pairs)** |  |  |  |  |  |
| Gamma | 5.528 (4.585-6.528) | 3.475 (2.590-4.265) | 5.323 (4.412-6.296) | 4.661 (3.482-5.706) | -277.32 |
| Lognormal | 5.521 (4.578-6.525) | 3.466 (2.593-4.263) | 5.220 (4.332-6.176) | 4.579 (3.454-5.583) | -275.71 |
| Weibull | 5.377 (4.450-6.421) | 4.303 (3.050-5.112) | 5.622 (4.698-6.672) | 5.882 (4.080-7.052) | -291.33 |
| **Only positive values of serial interval (51 pairs)** |  |  |  |  |  |
| Gamma | 5.880 (4.980-6.880) | 3.304 (2.437-4.150) | 5.274 (4.479-6.186) | 4.198 (3.152-5.190) | -246.03 |
| Lognormal | 5.967 (5.053-6.994) | 3.940 (2.719-5.302) | 4.980 (4.203-5.865) | 4.152 (3.129-5.126) | -247.6 |
| Weibull | 5.905 (4.992-6.917) | 3.328 (2.426-4.123) | 5.446 (4.624-6.409) | 4.561 (3.390-5.531) | -248.68 |

\*If the difference in AIC > 10 between two fits on same data, the difference in fit is significant with lower AIC suggesting a better fit.

**Table S4:**
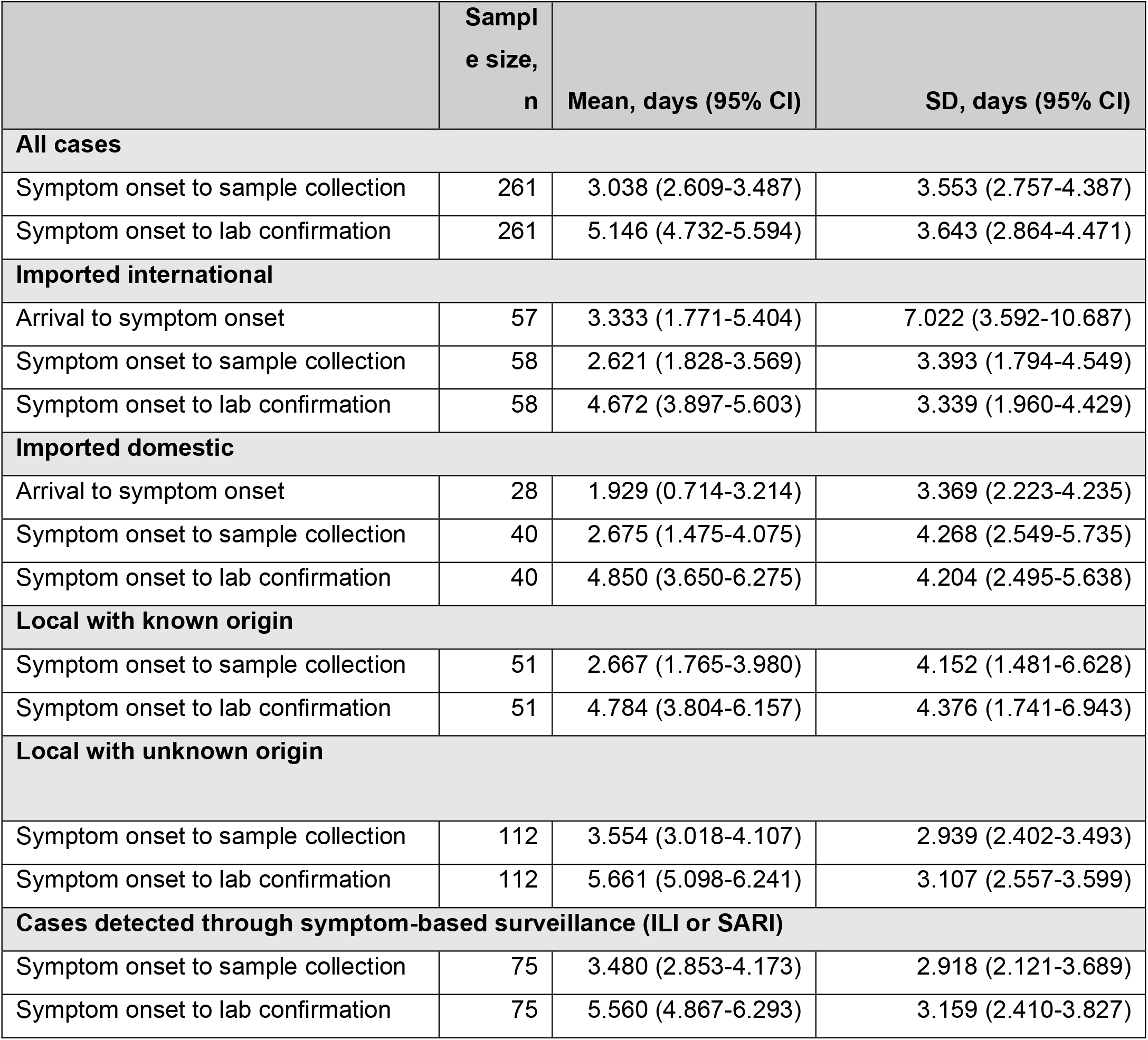
Estimates of various delays for COVID-19 cases up to 1 June 2020.

**Figure S1:**
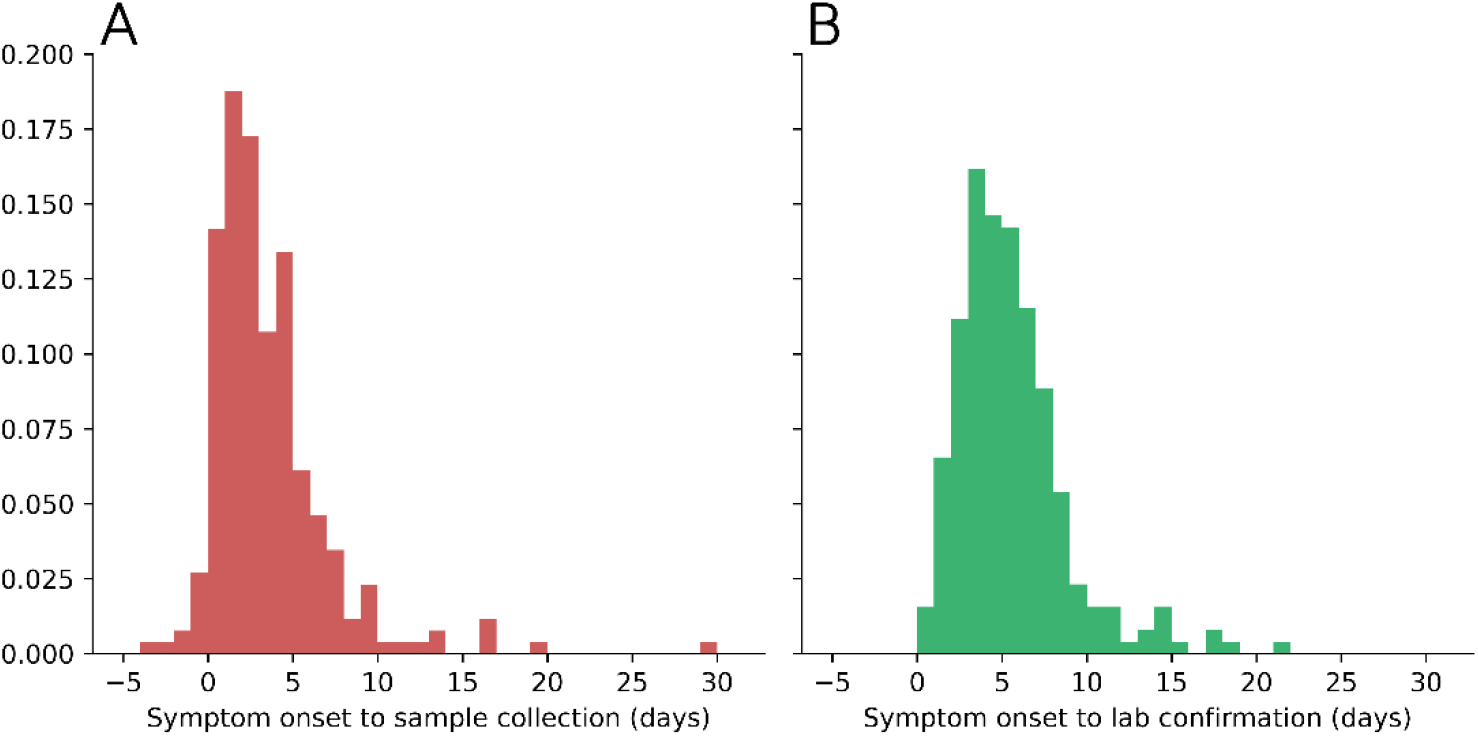
Histograms showing distributions of various delays for 261 symptomatic COVID-19 cases till 1 June 2020. **[A]** Delay from symptom onset to sample collection. **[B]** Delay from symptom onset to lab confirmation. See estimates in table S4.

**Figure S2:**
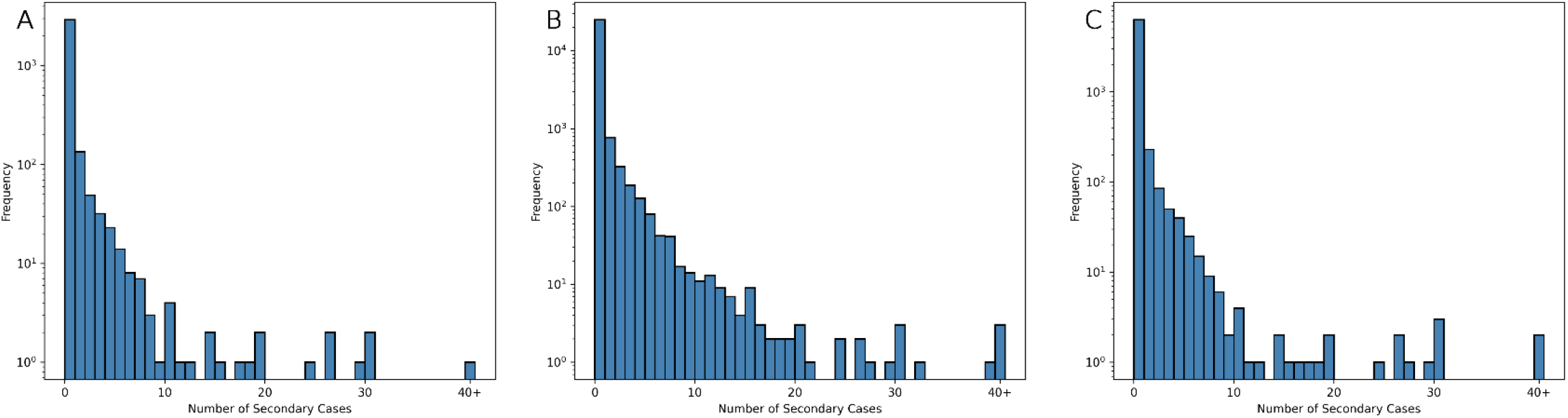
Observed offspring distribution of COVID-19 cases in Karnataka, India. Bars show observed frequency of the number of individuals infected by each case. **[A]** Till 1 June (n=3404), **[B]** Till 13 June (n=6824), **[C]** Till 21 July (n=71068). See estimates in table S2.

## References

1. The origin of SARS-CoV-2 - The Lancet Infectious Diseases [Internet]. [cited 2020 Oct 1]. Available from: https://www.thelancet.com/journals/laninf/article/PIIS1473-3099(20)30641-1/fulltext?rss=yes

2. COVID-19 Map - Johns Hopkins Coronavirus Resource Center [Internet]. [cited 2020 Sep 23]. Available from: https://coronavirus.jhu.edu/map.html

3. Karnataka Government [Internet]. [cited 2020 Nov 10]. Available from: https://karnataka.gov.in/info-1/State+Indicators/%E0%B2%9C%E0%B2%A8%E0%B2%B8%E0%B2%82%E0%B2%96%E0%B3%8D%E0%B2%AF%E0%B3%86/en

4. “A Menace to the Public Health” — Contact Tracing and the Limits of Persuasion \textbar NEJM [Internet]. [cited 2020 Sep 30]. Available from: https://www.nejm.org/doi/10.1056/NEJMp2021887?url_ver=Z39.88-2003&rfr_id=ori:rid:crossref.org&rfr_dat=cr_pub%20%200pubmed

5. Hellewell J, Abbott S, Gimma A, Bosse NI, Jarvis CI, Russell TW, et al. Feasibility of controlling COVID-19 outbreaks by isolation of cases and contacts. The Lancet Global Health. 2020 Apr 1;8(4):e488–96.

6. Superspreading and the effect of individual variation on disease emergence | Nature [Internet]. [cited 2020 Nov 28]. Available from: https://www.nature.com/articles/nature04153

7. Frieden TR, Lee CT. Identifying and Interrupting Superspreading Events—Implications for Control of Severe Acute Respiratory Syndrome Coronavirus 2 - Volume 26, Number 6—June 2020 - Emerging Infectious Diseases journal - CDC. [cited 2020 Nov 28]; Available from: https://wwwnc.cdc.gov/eid/article/26/6/20-0495_article

8. Woolhouse MEJ, Dye C, Etard J-F, Smith T, Charlwood JD, Garnett GP, et al. Heterogeneities in the transmission of infectious agents: Implications for the design of control programs. PNAS. 1997 Jan 7;94(1):338–42.

9. Vink MA, Bootsma MCJ, Wallinga J. Serial intervals of respiratory infectious diseases: a systematic review and analysis. Am J Epidemiol. 2014 Nov 1;180(9):865–75.

10. Rai B, Shukla A, Dwivedi LK. Estimates of serial interval for COVID-19: A systematic review and meta-analysis. Clin Epidemiol Glob Health. 2020 Aug 26;

11. Laxminarayan R, Wahl B, Dudala SR, Gopal K, B CM, Neelima S, et al. Epidemiology and transmission dynamics of COVID-19 in two Indian states. Science. 2020 Nov 6;370(6517):691–7.

12. Group ICS, Team CE & DM, Team CL, Team V. Laboratory surveillance for SARS-CoV-2 in India: Performance of testing & descriptive epidemiology of detected COVID-19, January 22 - April 30, 2020. Indian Journal of Medical Research. 2020 May 1;151(5):424.

13. Vasudevan V, Gnanasekaran A, Sankar V, Vasudevan SA, Zou J. Variation in COVID-19 Data Reporting Across India: 6 Months into the Pandemic. J Indian Inst Sci. 2020 Oct 1;100(4):885–92.

14. 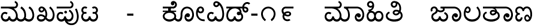 [Internet]. [cited 2020 Nov 17]. Available from: https://covid19.karnataka.gov.in/

15. India COVID-19 [Internet]. [cited 2020 Oct 3]. Available from: https://www.isibang.ac.in/~athreya/incovid19/data.html

16. STROBE_checklist_v4_combined.pdf [Internet]. [cited 2020 Nov 23]. Available from: https://www.equator-network.org/wp-content/uploads/2015/10/STROBE_checklist_v4_combined.pdf

17. Testing Strategy [Internet]. [cited 2020 Nov 17]. Available from: https://www.icmr.gov.in/cteststrat.html

18. Revised Quarantine and Testing Protocol for Primary-High Risk Contacts and Secondary - Low Risk Contacts.pdf [Internet]. [cited 2020 Nov 17]. Available from: https://covid19.karnataka.gov.in/storage/pdf-files/cir-hws/Revised%20Quarantine%20and%20Testing%20Protocol%20for%20Primary-High%20Risk%20Contacts%20and%20Secondary%20-%20Low%20Risk%20Contacts.pdf

19. 5543723831596613278.pdf [Internet]. [cited 2020 Nov 17]. Available from: https://ncdc.gov.in/WriteReadData/l892s/5543723831596613278.pdf

20. Guidelines for Inter State travelers to Karnataka during Phase Reopening (Unlock 1).pdf [Internet]. [cited 2020 Nov 17]. Available from: https://covid19.karnataka.gov.in/storage/pdf-files/FAQs/Guidelines%20for%20Inter%20State%20travelers%20to%20Karnataka%20during%20Phase%2 0Reopening%20(Unlock%201).pdf

21. Lr from OSD, DoHFW on sharing Best Practices in Contact tracing and household survey dated 18062020.pdf [Internet]. [cited 2020 Nov 17]. Available from: https://covid19.karnataka.gov.in/storage/pdf-files/Lr%20from%20OSD,%20DoHFW%20on%20sharing%20Best%20Practices%20in%20Contact%20tracing%20and%20household%20survey%20dated%2018062020.pdf

22. Considerations for implementing and adjusting public health and social measures in the context of COVID-19 [Internet]. [cited 2020 Dec 3]. Available from: https://www.who.int/publications-detail-redirect/considerations-in-adjusting-public-health-and-social-measures-in-the-context-of-covid-19-interim-guidance

23. Adam DC, Wu P, Wong JY, Lau EHY, Tsang TK, Cauchemez S, et al. Clustering and superspreading potential of SARS-CoV-2 infections in Hong Kong. Nature Medicine. 2020 Sep 17;1–6.

24. Modified Poisson Regression Approach to Prospective Studies with Binary Data | American Journal of Epidemiology | Oxford Academic [Internet]. [cited 2020 Nov 23]. Available from: https://academic.oup.com/aje/article/159/7/702/71883

25. epicontacts package | R Documentation [Internet]. [cited 2020 Nov 23]. Available from: https://www.rdocumentation.org/packages/epicontacts/versions/1.1.0

26. Buradikatti K. Ballari’s Jindal steel plant emerges COVID-19 cluster. The Hindu [Internet]. 2020 Jun 15 [cited 2020 Dec 3]; Available from: https://www.thehindu.com/news/national/karnataka/ballaris-jindal-steel-plant-emerges-covid-19-cluster/article31836343.ece

27. Bureau K. Pharma company case: A month on, source of infection remains mystery. The Hindu [Internet]. 2020 Apr 25 [cited 2020 Dec 3]; Available from: https://www.thehindu.com/news/national/karnataka/govt-orders-probe-into-spread-of-covid-19-from-nanjangud-pharma-company/article31431712.ece

28. Althouse BM, Wenger EA, Miller JC, Scarpino SV, Allard A, Hébert-Dufresne L, et al. Superspreading events in the transmission dynamics of SARS-CoV-2: Opportunities for interventions and control. PLOS Biology. 2020 Nov 12;18(11):e3000897.

29. Kim Y, Ryu H, Lee S. Agent-Based Modeling for Super-Spreading Events: A Case Study of MERS-CoV Transmission Dynamics in the Republic of Korea. International Journal of Environmental Research and Public Health. 2018 Nov;15(11):2369.

30. Goyal A, Reeves DB, Cardozo-Ojeda EF, Schiffer JT, Mayer BT. Wrong person, place and time: viral load and contact network structure predict SARS-CoV-2 transmission and super-spreading events. medRxiv [Internet]. 2020 Sep 28 [cited 2020 Nov 30]; Available from: https://www.ncbi.nlm.nih.gov/pmc/articles/PMC7536880/

31. Endo A, Centre for the Mathematical Modelling of Infectious Diseases COVID-19 Working Group, Abbott S, Kucharski AJ, Funk S. Estimating the overdispersion in COVID-19 transmission using outbreak sizes outside China. Wellcome Open Res. 2020 Jul 10;5:67.

32. Hasan A, Susanto H, Kasim M, Nuraini N, Lestari B, Triany D, et al. Superspreading in Early Transmissions of COVID-19 in Indonesia [Internet]. Epidemiology; 2020 Jun [cited 2020 Nov 30]. Available from: http://medrxiv.org/lookup/doi/10.1101/2020.06.28.20142133

33. Zhang Y, Li Y, Wang L, Li M, Zhou X. Evaluating Transmission Heterogeneity and Super-Spreading Event of COVID-19 in a Metropolis of China. International Journal of Environmental Research and Public Health. 2020 Jan;17(10):3705.

34. Bi Q, Wu Y, Mei S, Ye C, Zou X, Zhang Z, et al. Epidemiology and transmission of COVID-19 in 391 cases and 1286 of their close contacts in Shenzhen, China: a retrospective cohort study. The Lancet Infectious Diseases. 2020 Aug;20(8):911–9.

35. Lau MSY, Grenfell B, Thomas M, Bryan M, Nelson K, Lopman B. Characterizing superspreading events and age-specific infectiousness of SARS-CoV-2 transmission in Georgia, USA. PNAS. 2020 Sep 8;117(36):22430–5.

36. Lemieux J, Siddle KJ, Shaw BM, Loreth C, Schaffner S, Gladden-Young A, et al. Phylogenetic analysis of SARS-CoV-2 in the Boston area highlights the role of recurrent importation and superspreading events. medRxiv. 2020 Aug 25;2020.08.23.20178236.

37. Kretzschmar ME, Rozhnova G, Bootsma MCJ, Boven M van, Wijgert JHHM van de, Bonten MJM. Impact of delays on effectiveness of contact tracing strategies for COVID-19: a modelling study. The Lancet Public Health. 2020 Aug;5(8):e452–e459.

38. Luo L, Liu D, Liao X, Wu X, Jing Q, Zheng J, et al. Contact Settings and Risk for Transmission in 3410 Close Contacts of Patients With COVID-19 in Guangzhou, China. Ann Intern Med [Internet]. 2020 Aug 13 [cited 2020 Nov 10]; Available from: https://www.ncbi.nlm.nih.gov/pmc/articles/PMC7444623/

39. Davies NG, Klepac P, Liu Y, Prem K, Jit M, Eggo RM. Age-dependent effects in the transmission and control of COVID-19 epidemics. Nature Medicine. 2020 Aug;26(8):1205–11.

40. He D, Zhao S, Lin Q, Zhuang Z, Cao P, Wang MH, et al. The relative transmissibility of asymptomatic COVID-19 infections among close contacts. International Journal of Infectious Diseases. 2020 May;94:145–7.

41. SARS-CoV-2 viral load is associated with increased disease severity and mortality | Nature Communications [Internet]. [cited 2020 Dec 3]. Available from: https://www.nature.com/articles/s41467-020-19057-5

42. Beldomenico PM. Do superspreaders generate new superspreaders? A hypothesis to explain the propagation pattern of COVID-19. International Journal of Infectious Diseases. 2020 Jul 1;96:461–3.

43. Saurabh S, Verma MK, Gautam V, Kumar N, Goel AD, Gupta MK, et al. Transmission Dynamics of the COVID-19 Epidemic at the District Level in India: Prospective Observational Study. JMIR Public Health and Surveillance. 2020;6(4):e22678.

44. Ali ST, Wang L, Lau EHY, Xu X-K, Du Z, Wu Y, et al. Serial interval of SARS-CoV-2 was shortened over time by nonpharmaceutical interventions. Science. 2020 Aug 28;369(6507):1106–9.

45. Wallinga J, Lipsitch M. How generation intervals shape the relationship between growth rates and reproductive numbers. Proc Biol Sci. 2007 Feb 22;274(1609):599–604.

46. Al-Qahtani M, AlAli S, Rahman AKA, Alsayyad AS, Otoom S, Atkin SL. The prevalence of asymptomatic and symptomatic COVID19 disease in a cohort of quarantined subjects. International Journal of Infectious Diseases [Internet]. 2020 Nov 2 [cited 2020 Nov 23];0(0). Available from: https://www.ijidonline.com/article/S1201-9712(20)32294-3/abstract

47. He J, Guo Y, Mao R, Zhang J. Proportion of asymptomatic coronavirus disease 2019: A systematic review and meta-analysis. Journal of Medical Virology [Internet]. [cited 2020 Nov 23];n/a(n/a). Available from: https://onlinelibrary.wiley.com/doi/abs/10.1002/jmv.26326

48. Liu Z, Chu R, Gong L, Su B, Wu J. The assessment of transmission efficiency and latent infection period in asymptomatic carriers of SARS-CoV-2 infection. International Journal of Infectious Diseases. 2020 Oct;99:325– 7.

49. Buitrago-Garcia D, Egli-Gany D, Counotte MJ, Hossmann S, Imeri H, Ipekci AM, et al. Occurrence and transmission potential of asymptomatic and presymptomatic SARS-CoV-2 infections: A living systematic review and meta-analysis. PLOS Medicine. 2020 Sep 22;17(9):e1003346.

## References (for supplementary materials)

1 Health and Family Welfare Department, Government of Karnataka. Revised Quarantine and Testing Protocol for Primary-High Risk Contacts and Secondary - Low Risk Contacts. https://covid19.karnataka.gov.in/storage/pdf-files/cir-hws/Revised%20Quarantine%20and%20Testing%20Protocol%20for%20Primary-High%20Risk%20Contacts%20and%20Secondary%20-%20Low%20Risk%20Contacts.pdf.

2 Li R, Pei S, Chen B, et al. Substantial undocumented infection facilitates the rapid dissemination of novel coronavirus (SARS-CoV2). Science 2020; published online March 16. DOI:10.1126/science.abb3221.

3 Adam DC, Wu P, Wong JY, et al. Clustering and superspreading potential of SARS-CoV-2 infections in Hong Kong. Nat Med 2020; 26: 1714–9.

4 Lau MSY, Grenfell B, Thomas M, Bryan M, Nelson K, Lopman B. Characterizing superspreading events and age-specific infectiousness of SARS-CoV-2 transmission in Georgia, USA. Proc Natl Acad Sci 2020; 117: 22430–5.

5 Knight G, Leclerc QJ, Kucharski AJ. Analysis of SARS-CoV-2 transmission clusters and superspreading events. 2020; published online June 3.

6 Zou G. A Modified Poisson Regression Approach to Prospective Studies with Binary Data. Am J Epidemiol 2004; 159: 702–6.

7 Public health criteria to adjust public health and social measures in the context of COVID-19: annex to considerations in adjusting public health and social measures in the context of COVID-19, 12 May 2020. World Health Organization, 2020.

8 Considerations in adjusting public health and social measures in the context of COVID-19: interim guidance, 16 April 2020. World Health Organization, 2020.

9 Indian Council of Medical Research (ICMR). Testing Strategy Updates for COVID-19 in India. https://www.icmr.gov.in/cteststrat.html (accessed Dec 3, 2020).

